# Exploring the relationship between job characteristics and infection: Application of a COVID-19 Job Exposure Matrix to SARS-CoV-2 infection data in the United Kingdom

**DOI:** 10.1101/2022.09.21.22280191

**Authors:** Sarah Rhodes, Sarah Beale, Jack Wilkinson, Karin van Veldhoven, Ioannis Basinas, William Mueller, Karen Oude Henge, Alex Burdorf, Susan Peters, Zara A Stokholm, Vivi Schlünssen, Henrik Kolstad, Anjoeka Pronk, Neil Pearce, Andrew Hayward, Martie van Tongeren

## Abstract

**Objectives:** To assess whether workplace exposures as estimated via a COVID-19 Job Exposure Matrix (JEM) are associated with SARS-CoV-2.

**Methods:** Data on 244,470 participants were available from the ONS Coronavirus Infection Survey (CIS) and 16,801 participants from the Virus Watch Cohort, restricted to workers aged 20 to 64. Analysis used logistic regression models with SARS-CoV-2 as the dependent variable for eight individual JEM domains (number of workers, nature of contacts, contact via surfaces, indoor or outdoor location, ability to social distance, use of face covering, job insecurity, migrant workers) with adjustment for age, sex, ethnicity, Index of Multiple Deprivation (IMD), region, household size, urban vs rural area, and health conditions. Analyses were repeated for three time periods (i) February 2020 (Virus Watch)/April 2020 (CIS) to May 2021), (ii)June 2021 to November 2021, (iii) December 2021 to January 2022.

**Results:** Overall, higher risk classifications for the first six domains tended to be associated with an increased risk of infection, with little evidence of a relationship for domains relating to proportion of workers with job insecurity or migrant workers. By time there was a clear exposure-response relationship for these domains in the first period only. Results were largely consistent across the two cohorts.

**Conclusions:** An exposure-response relationship exists in the early phase of the COVID-19 pandemic for number of contacts, nature of contacts, contacts via surfaces, indoor or outdoor location, ability to social distance and use of face coverings. These associations appear to have diminished over time.

## Background

COVID-19 has been responsible for millions of deaths globally (1). Risk of infection with SARS-CoV-2 has been found to vary with occupation (2, 3), and differences in workplace exposure is likely to explain some of this variation (4, 5). Accordingly, there is interest in identifying the occupations at increased risk and workplace features and mitigation strategies which modulate this risk. Workplace factors associated with exposure include number and type of daily contacts, ability to socially distance, and workplace ventilation (4). Studies of social contact patterns (6, 7) suggest that workers in retail, hospitality, healthcare, education and transportation have the highest number of workplace contacts. Baker et al. (8) identified healthcare, protective service, office and administrative support, education, community and social services and construction and extraction occupations as all having a high level of exposure to infection. Evidence is varied on whether this perceived exposure translates to increased COVID-19 disease and mortality (3, 9-11).

The COVID-19 Job Exposure Matrix (JEM), has been developed to categorise occupations according to workplace factors believed to be associated with SARS-CoV-2 infection (4). This JEM was based on assessment from occupational exposure experts from three countries (Denmark, the Netherlands, United Kingdom) regarding eight distinct risk domains related to the risk of transmission ((1) number of contacts, (2) nature of contacts, (3) contaminated workspaces, (4) location of the worksite)), the presence of mitigation measures ((5) social distance and (6) the use of face covering) and the level of precarity of the occupation involved ((7) proportion of workers with income insecurity and (8) proportion of migrant workers). Distinct JEM scores for each country were established to account for the presence of different guidelines and mitigation measures at a time following the first lockdown. A validation exercise using 6,794 participants has already been performed in the Netherlands involving comparison of the risk scores assigned by the COVID-19-JEM against self-reported data from surveys performed for this purpose (12). Results suggested good agreement between the JEM scores and the self-reported data for most dimensions except face covering. Validation of the separate JEM domains against self-reported COVID-19 illness was also performed. Higher COVID-19-JEM assigned risk scores were associated with higher odds ratios (range: 1.28-1.80) of COVID-19 for all except the precarious dimensions (12).

In the present study we implemented the UK specific risk scores/assignment of the COVID-19-JEM on two separate large UK studies containing information on occupation and SARS-CoV-2 infections with the aims to a) evaluate the performance of the UK edition of the COVID-19 JEM as a tool for assessing occupational exposure to SARS-CoV-2 b) assess the relationship between the exposure affecting factors included in the JEM and SARS-CoV-2 infection risk and (c) assess whether this relationship is consistent over time.

### Description of JEM

The COVID-19 Job Exposure Matrix (JEM) was established using expert judgement and consensus (4). The JEM was developed with reference to the conditions present during the period following the easing of the first strict lockdown measures. It was anticipated that workers were encouraged to work from home, where possible, but those who needed to attend the workplace were allowed to work. It was also assumed that hand washing, use of personal protective equipment (PPE) and face coverings and social distancing in the workplace were advised, and vaccination programmes had not yet started. The JEM contained eight domains representing factors which were judged to affect occupational exposure to SARS-CoV-2, classified by 4 risk levels (Table 1). The COVID-19 JEM was coded according to the International Standard Classification of Occupations 2008 (ISCO-08) coding system (13) and includes specific scores for the Netherlands, Denmark and the UK.

**Table 1.** Description of 8 domains of the Job Exposure Matrix (JEM)

| **Dimension** | **Description** | **Abbreviation used in figures** | **Levels** |
| --- | --- | --- | --- |
| **1** | **The number of workers at worksite** | **Number** | **Homeworking/lone working**<br>*(No risk)*<br>**<10 workers/day** *(Low risk)*<br>**10-30 workers/day** *(Elevated risk)*<br>**>30 workers/day** *(High risk)* |
| **2** | **The nature of contacts with co-workers, general public or patients with COVID-19** | **Nature** | **Homeworking/lone working**<br>*(No risk)*<br>**co-workers only**<br>*(Low risk)*<br>**General public**<br>*(Elevated risk)*<br>**Patients, including with C19**<br>*(High risk)* |
| **3** | **The risk through contaminated work surfaces and materials** | **Surfaces** | **Homeworking/lone working**<br>*(No risk)*<br>**Frequently sharing contact surfaces with co-workers**<br>*(Low risk)*<br>**Occasionally sharing contact surfaces with general public**<br>*(Elevated risk)*<br>**Frequently sharing contact surfaces with general public**<br>*(High risk)* |
| **4** | **Location of work: indoors or outdoors** | **Location** | **Homeworking/lone working**<br>*(No risk)*<br>**Mostly outdoors** *(Low risk)*<br>**Partly indoor** *(Elevated risk)* |
|  |  |  | **Mostly indoor (*High risk*)** |
| **5** | **The possibility to keep at least 1m of social distance** | **Distancing** | **Homeworking/lone working (*No risk*)**<br>**Always maintained (*Low risk*)**<br>**Cannot always be maintained (*Elevated risk*)**<br>**can never be maintained (*High risk*)** |
| **6** | **The need and usage of face covering** | **Face covering** | **Homeworking/lone working (*No risk*)**<br>**Always (*Low risk*)**<br>**Not always while in proximity to others (*Elevated risk*)**<br>**Face covering not feasible (*High risk*)** |
| **7** | **Job insecurity: proportion of flexible labour contracts** | **Insecurity** | **0 (*None*)**<br>**1-10% (*Low*)**<br>**11-25% (*Elevated*)**<br>**>25% (*High*)** |
| **8** | **Migrant workers: proportion of migrant workers** | **Migrants** | **0 (*None*)**<br>**1-10% (*Low*)**<br>**11-25% (*Elevated*)**<br>**>25% (*High*)** |

For the UK component the COVID-19 JEM was translated from the ISCO-08 coding system to the UK Standard Occupational Classification (SOC) 2010 version coding system (14).Translation was facilitated by a crosswalk developed by the UK Office for National Statistics (ONS) (15). Evaluation of occupation descriptions between the two coding systems revealed that there was not direct correspondence with 46 ISCO codes not represented in the SOC system and 42 SOC codes not represented in the ISCO system. For those the domains of the JEM related to risk for transmission and mitigation measures were scored by the same three UK experts involved in the development of the main JEM using the same consensus procedure. Domains related to income insecurity and migrant workers used UK data extracted from the GB and Scottish components of the Annual Population Survey (APS) April 2019 to March 2020 (16) and the UK broad Labour Force Survey (LFS) August to October 2010 (17), respectively (as utilised in the original JEM). To minimise bias due to changes in perception of experts by time, the translation to SOC 2010 occurred simultaneously with the original JEM.

### Datasets

Virus Watch is a prospective household cohort study based in England and Wales (*n*=58,560 participants from 28,449 households). Participants provide demographic and health-related information at enrolment; they then complete weekly surveys reporting any symptoms, test results of lateral flow or PCR tests taken under national testing scheme, and vaccinations, as well as monthly surveys concerning socio-behavioural and clinical factors. Subsets of participants, received in-clinic serological testing, and/or performed monthly at-home finger-prick serological tests. All participants’ records were linked to national databases of SARS-CoV-2 PCR and lateral flow test results. SARS-CoV-2 infection status relates to any evidence of infection via self-reported, national database or serological test. Upon study enrolment, participants are asked about their employment status and, if employed or self-employed, prompted to enter free text for their job title; four-digit SOC codes were derived from free-text job titles using semi-automatic processing using Cascot Version 5.6.3 (https://warwick.ac.uk/fac/soc/ier/software/cascot/) – the ONS-recommended methodology for assigning SOC codes to free-text data. The survey began in June 2020 but retrospectively looked at infection status from Feb 2020 onwards. Data used relates to Feb 2020 to Jan 2022. Further details on the Virus Watch cohort and methodology can be found on its study protocol available at https://bmjopen.bmj.com/content/11/6/e048042.

The ONS COVID-19 Infection survey (CIS) is a repeated cross-sectional household survey designed to be representative of the UK population for calculation of monthly UK prevalence estimates of SARS-CoV-2 virus. Participation starts with five weeks of weekly visits for each household, followed by monthly visits. Each visit includes a survey and a Covid-19 PCR test for each household member regardless of symptoms. At each visit, participants are asked about their work status and job title; free text job titles were used by the ONS to derive four-digit SOC codes via a combination of automatic methods and manual coding. We used the data from April 2020 to Jan 2022 which was accessed via the Secure Research Server (SRS) using Stata 17 (18). A detailed description of the CIS methodology is provided within its study protocol available at https://www.ndm.ox.ac.uk/covid-19/covid-19-infection-survey/protocol-and-information-sheets

### Statistical methods

All analyses were restricted to working age adults (20-64 years) who reported being employed or self-employed at enrolment into the studies. Missing covariate data were known to be sparse across adjustment covariates (0-1.3%) so we restricted data to complete cases on these variables. Relationships between the individual JEM risk domains and SARS-CoV-2 infection status (ever/never within the time period of interest) were evaluated using logistic regression. In Virus Watch, the infection status was derived based on any serological or virological evidence of infection (positive lateral flow (LFT), polymerase chain reaction (PCR), anti-nucleocapsid antibody serological test, or anti-spike antibody serological test in absence of vaccination), whereas in CIS only PCR tests conducted as part of the survey were considered.

Four digit SOC codes were used to derive covariates relating to perceived occupational exposure on the 8 JEM domains. These related to the first available SOC within each time period for CIS and baseline occupation for Virus Watch. ‘No risk’ was set as the reference category for all JEM exposures, with the exception of Domain 7 (migrant workers) for Virus Watch where it was set at ‘low risk’ due to small cell sizes in the ‘no risk’ category. A ‘missing’ category was included for working participants for whom a four digit SOC code was not available and this was used in summary tables and regression (for comparison). The coefficients relating to the missing category are not presented in the coefficient plots. A correlation matrix of Spearmans rank correlation between derived JEM domain scores was produced.

Potential confounders were selected and included on the models based on a directed acylic graphs (DAG) an interactive version of which is available at http://dagitty.net/dags.html?id=mGNoZU. We adjusted all estimates for age (5 ordered categories), sex, minority ethnicity (White British versus other), geographic region (ONS national region), deprivation based on Indices of Multiple Deprivation (IMD) derived from postcode), household size, urban versus rural area, and health status. For Virus Watch clinically vulnerable (yes/no) was defined as any condition on the UK NHS/government list of clinically vulnerable conditions, obesity, and/or having received an NHS shielding letter. For the CIS, health conditions was a yes response to the question ‘Do you have any physical or mental health conditions or illnesses lasting or expected to last 12 months or more (excluding any long-lasting COVID-19 symptoms)’?

To investigate whether the association between JEM domains and risk of infection varied across different pandemic phases, we then repeated the analysis stratified by time period. The first time period corresponded to the first and second waves of the pandemic (up to May 2021), the second to the third pandemic wave (June 2021 to November 2021), and the third to the Omicron-dominated fourth wave (December 2021 - January 2022). For Virus Watch, Period 1 comprised a large amalgamated time period as serological testing began during the second wave, and thus infections could be attributed to either the first or second wave; mass population testing also only became available post-first-wave in England and Wales.

Infections that were derived from serological testing without a prior negative result could not be attributed to a particular date. JEM Domains relating to job insecurity and migrant workers were excluded from the time-stratified analyses due to low cells sizes for some risk levels.

For Virus Watch, to address differences in access and virological/antigen testing protocols across occupations, as well as address asymptomatic infection, we also performed a sensitivity analysis limited to participants who had undergone serological testing (*n*=6712). As above, JEM Domains relating to job insecurity and migrant workers were excluded from this analysis due to low cells sizes for some risk levels.

## Results

Demographic features of included Virus Watch and CIS participants are reported in Table 2. There were 244,470 participants from the CIS cohort and 16,479 from Virus Watch who met our inclusion criteria and were included in the analyses. Overall, the two studies shared comparable demographic distributions regarding gender and ethnicity. Virus watch contained a higher proportion of participants aged between 60 and 64 and in 1-2 person households with a greater proportion of participants residing in East England. Scotland was geographically represented only in the CIS study. Table 3 reports SARS-CoV-2 infection status by each risk category for all JEM domains for the full cohorts (CIS and Virus Watch) and serological sub-cohort (Virus Watch).There was strong correlation between some domains of the JEM, with the strongest correlations observed between the domains related to nature and surfaces (0.90) and number and nature (0.84) (S3).

**Table 2.** Demographic Characteristics of workers participating in the CIS and Virus Watch

| **Characteristic** | **CIS**<br>**N=244,470**<br>**n(%)** | **Virus Watch**<br>**N=16,479**<br>**n(%)** |
| --- | --- | --- |
| **Age** |  |  |
| 20-29 | 34,744(14%) | 1,560 (9.5%) |
| 30-39 | 55,436(22.7%) | 3,010 (18%) |
| 40-49 | 63,688(26%) | 3,661 (22%) |
| 50-59 | 67,803(28%) | 4,534 (28%) |
| 60-64 | 22,799(9%) | 3,714 (23%) |
| **Sex** |  |  |
| Female | 128,359(52%) | 9,090 (55%) |
| Male | 116,111(47%) | 7,389 (45%) |
| **Ethnic group** |  |  |
| White | 233,306(91%) | 14,687 (89%) |
| Black | 2,827(1.2%) | 208 (1.3%) |
| Mixed | 3,748(1.5%) | 308 (1.9%) |
| Asian | 11,837(4.8%) | 1,165 (7.1%) |
| Other Ethnicity | 2,605(1.0%) | 111 (0.7%) |
| **Health conditions** | 37,907(15.5%) |  |
| **Clinically vulnerable** |  | 8,606 (52%) |
| **IMD quartile (CIS)/quintine (VW)** |  |  |
| 1 | 36,376(15%) | 1,799 (11%) |
| 2 | 58,459(24%) | 2,945 (18%) |
| 3 | 70,300(29%) | 3,395 (21%) |
| 4 | 79,335(32%) | 4,057 (25%) |
|  |  | 4,283 (26%) |
| **Household Size** |  |  |
| 1 person | 31,977(13%) | 3,153 (19%) |
| 2 people | 89,782(37%) | 7,084 (43%) |
| 3 people | 51,003(21%) | 2,772 (17%) |
| 4 people | 51,521(21%) | 2,584 (16%) |
| 5 or more people | 20,187(8%) | 886 (5%) |
| **Region** |  |  |
| East Midlands | 15,239(6%) | 1,433 (8.7%) |
| East of England | 22,259(9%) | 3,311 (20%) |
| London | 51,554(21%) | 3,304 (20%) |
| North East | 8,092(3%) | 757 (4.6%) |
| North West | 26,830(11%) | 1,641 (10.0%) |
| South East | 30,117(12%) | 3,125 (19%) |
| South West | 18,252(7%) | 1,043 (6.3%) |
| Wales | 11,229(5%) | 340 (2.1%) |
| West Midlands | 17,312(7%) | 750 (4.6%) |
| Yorkshire and The Humber | 18,667(8%) | 775 (4.7%) |
| Scotland | 18,550(8%) | **NA** |

**Table 3.**
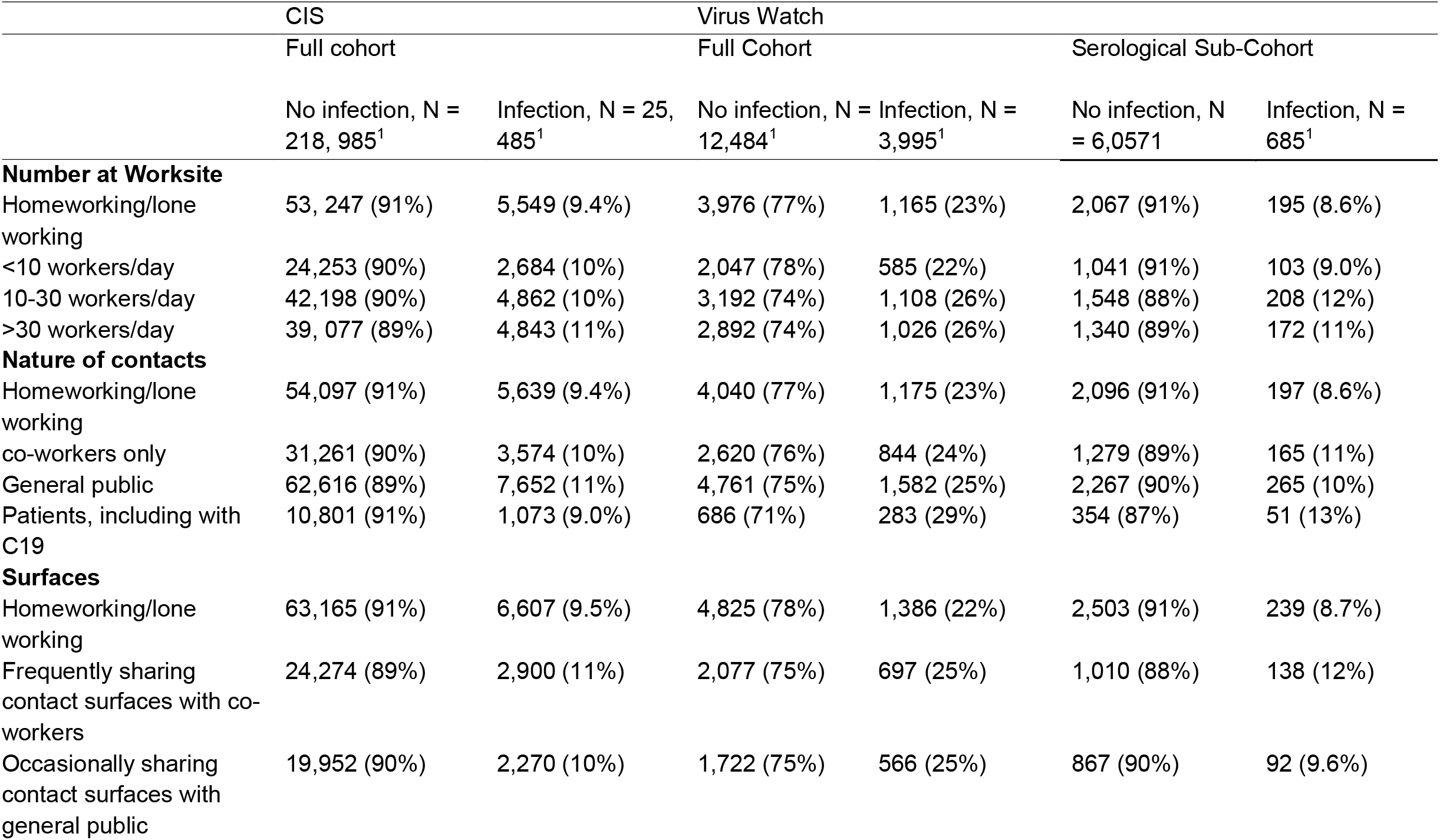
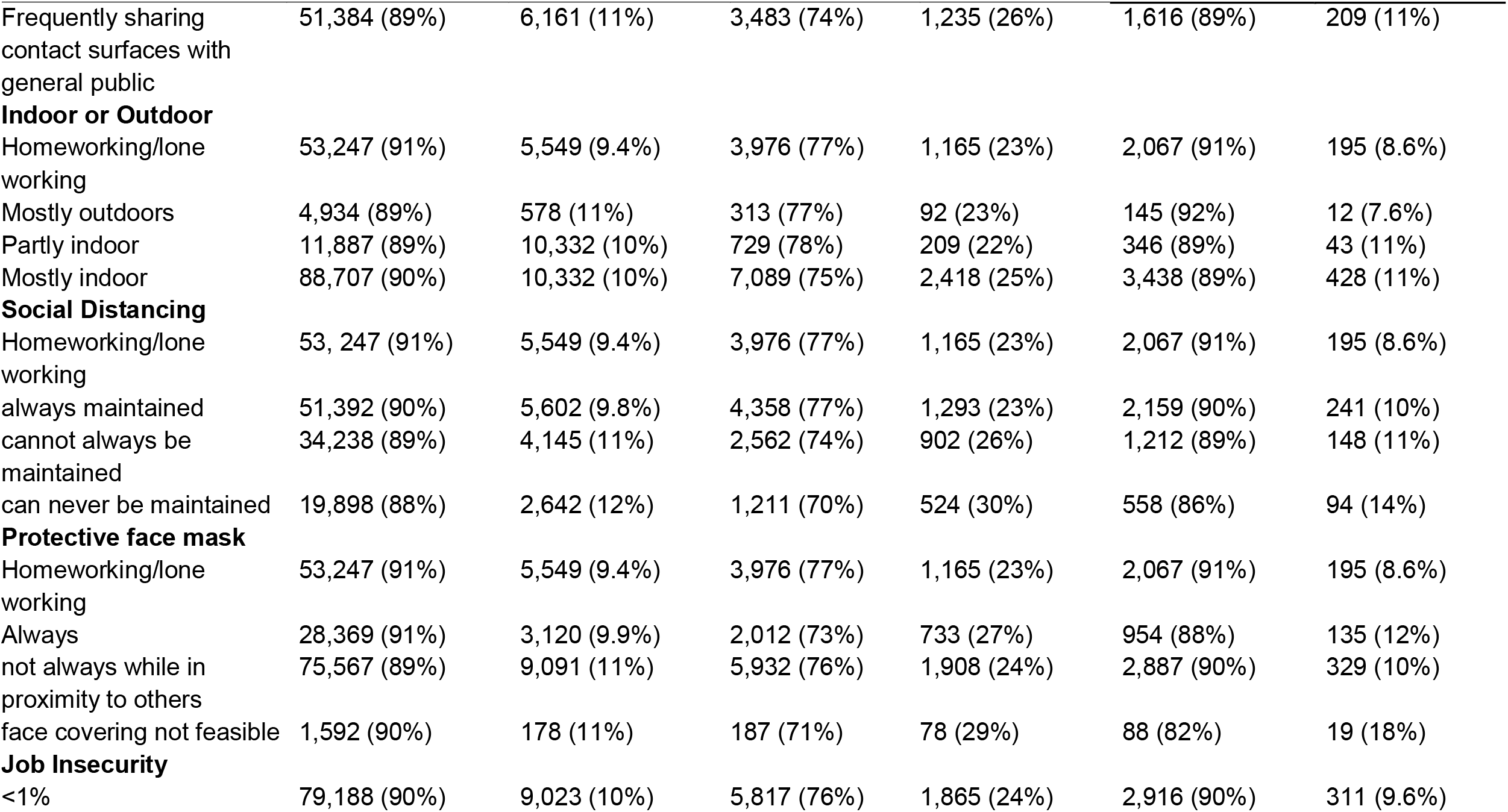
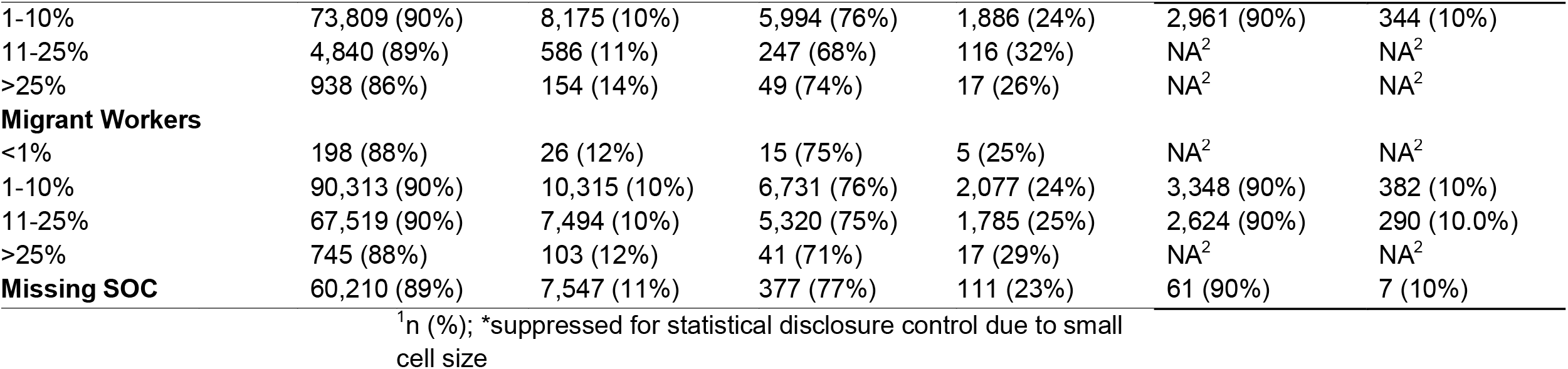
Participants with at least one infection by JEM domain by risk level for CIS and two Virus Watch cohorts (full cohort and serological sub-cohort).

Figure 1 shows the relationship between the first 6 JEM domains and COVID-19 infection. Estimates for all 8 domains are shown in S Table 1 and S Table 2, showing consistency across the two cohorts. Higher risk classifications were generally associated with increased risk of infection for the first 6 domains, although not always displaying an exposure-response relationship across all four categories. For domains relating to insecurity and migration, the association was less clear, such that classification as ‘No risk’ was not consistently lower than other risk categories (S2).

**Figure 1.**
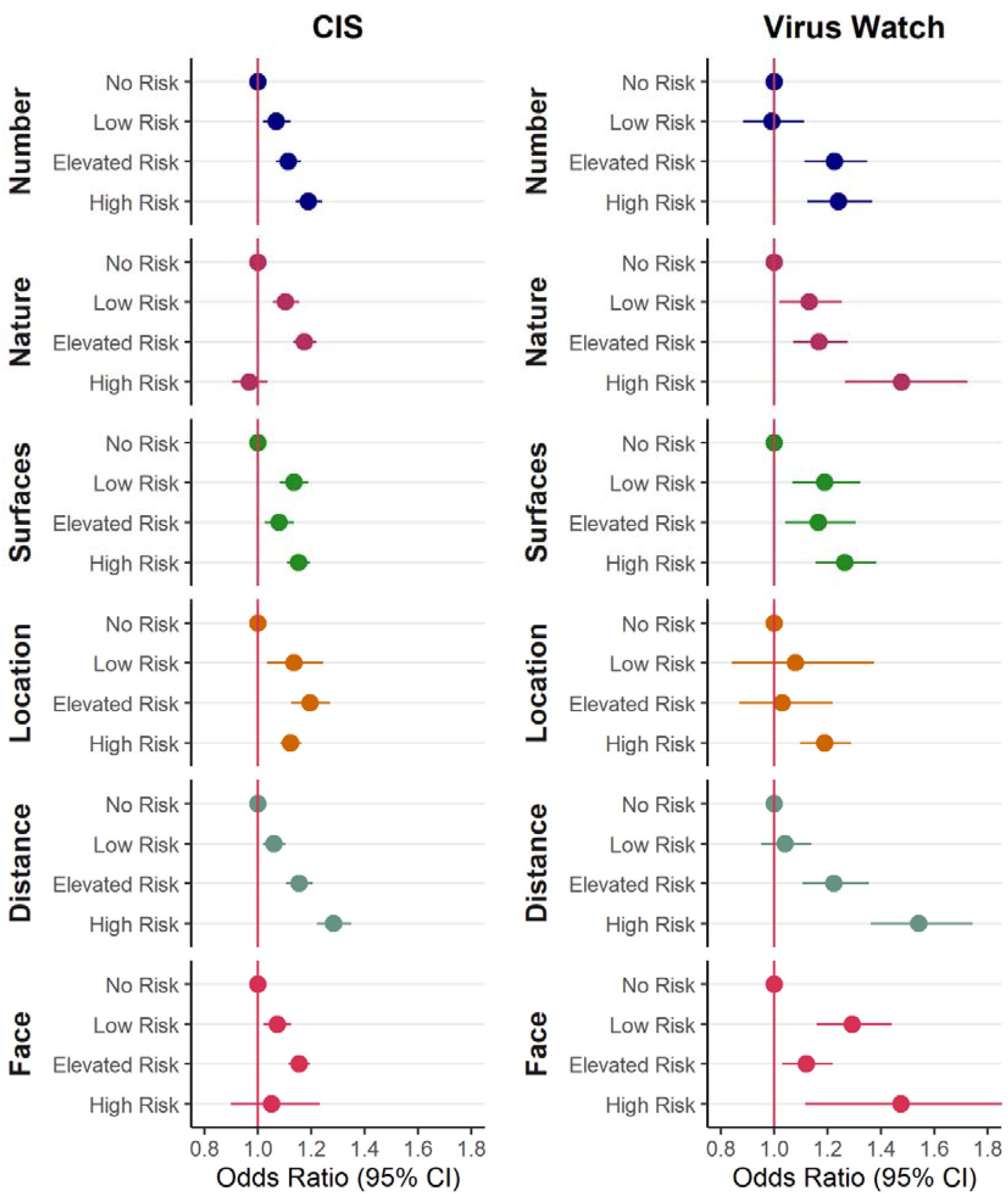
Odds ratios by level of exposure for 6 domains of the Job Exposure Matrix (compared to the no risk group) from (Virus Watch N= 16 801 and CISN=244 470) participants in work and aged 20-64. Models adjusted for age quintile, sex, ethnicity, IMD/health conditions, region, household size, urban vs rural area, and presence of health conditions. Domains relating to insecurity and migrant workers not displayed here due to imprecision. See Table 1 for explanation of abbreviations.

Figure 2 and Table 3 show the relationship between the domains of the JEM and COVID-19 infection during 3 time tranches. Generally, patterns across both cohorts and the Virus Watch serological sub-cohort are consistent. During time period 1, there is a clear exposure -response relationship in both cohorts between the level of risk attributed by the JEM and the risk of SARS-CoV-2 infection for the domains relating to the number of contacts, the nature of contacts and social distancing; with increasing odds ratios comparing low, elevated and high risk groups to the no risk group. Other domains (relating to surfaces, location and distancing) have odds ratios in the expected direction, showing increased risks for those in the low, elevated and high risk groups when compared to no risk, but the dose-response relationship is not evident. During time periods 2 and 3, there is little evidence of a relationship between the JEM domains and the risk of infection, most confidence intervals straddle the line of no difference and overlap. One exception is the domain relating to the nature of contacts; the ‘high risk’ group (contact with patients, including those suspected with COVID-19) in this domain is observed to have a reduced risk of infection in time periods 2 and 3 for both cohorts. Another exception is the domain relating to use of face coverings, where the low risk group appears to have a reduced risk of infection compared to no risk in time tranche 3 for the CIS data.

**Figure 2.**
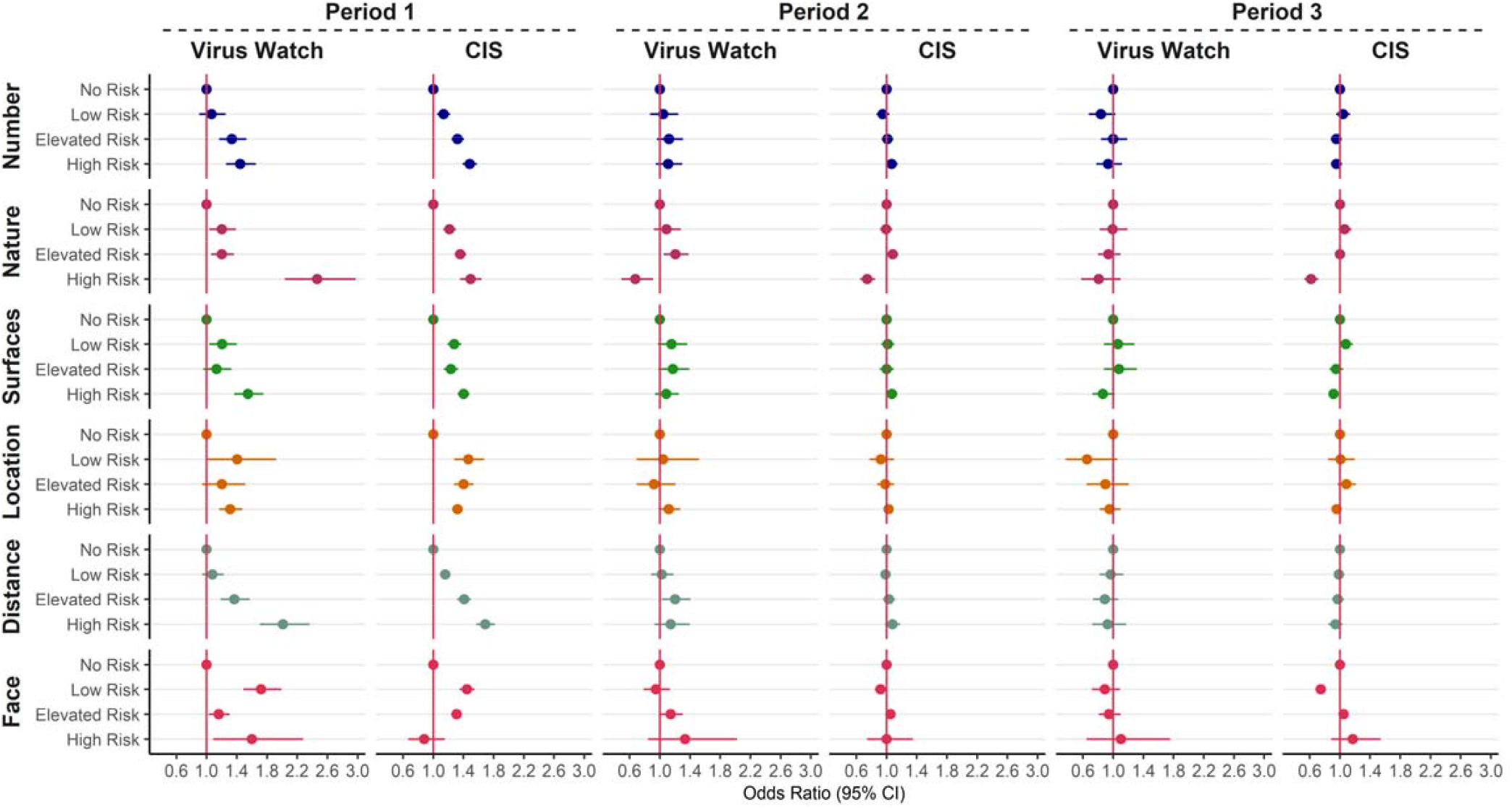
Odds ratios for COVID-19 by level of exposure for 6 domains of the Job Exposure Matrix (compared to the no risk group) from (Virus Watch N= 16 801 and CIS N=244 470) participants in work and aged 20-64. Data split into 3 time periods and models adjusted for age, sex, ethnicity, IMD, region, household size, urban vs rural area, and presence of health conditions. See Table 1 for explanation of abbreviations.

## Discussion

The present study evaluated the performance of a COVID-19 JEM by examining the relationships between its domains comprising occupational factors thought to be related to the risk of SARS-CoV-2 infection in two independent UK cohorts. Over the entire study period, a higher JEM score was associated with a higher risk of SARS-CoV-2 infection for the four domains of transmission risk (number of contacts, nature of contacts, contaminated work spaces, location) and the two domains of mitigation measures (social distancing, use of face covering) across both cohorts. However, after stratification by time period, this trend was evident only during the first time period across both cohorts.

Three domains (number, nature, and distancing) showed an exposure-response relationship between the risk level of the JEM and the risk of SARS-CoV-2 infection in the overall analysis. The absence of a clear relationships for the other domains could, at least partly, reflect the low number of participants working (mostly) outside (low risk), never using face covering (high risk) or having contact with the public (elevated risk), which can also explain the very broad confidence intervals estimated for those categories.

Interestingly, when stratified by time period the same associations were not observed in time period 2 or 3 in either cohort, which could be due to several reasons. Firstly the JEM was not updated to reflect different mitigation strategies in each time period; there is likely to be misclassification of exposure in those periods, especially with regards to the “home working / lone working” (no risk) category where large changes took place for a number of occupations following the implementation of a “return to work” policy in June 2021. Considering this group is the reference category this would likely result in bringing relationships towards the null. This was observed, particularly within the CIS, where the increasing trends between the low, medium and high risk categories remain for some dimensions, but not relative to the reference category (Figure 2). The JEM was developed on the knowledge from the first wave, when the theory of contaminated work surfaces was very prominent, however, with later evidence it has become clear that airborne transmission and ventilation was far more important (19), which we suggest should be reflected in updates of the JEM.

Secondly, a substantial part of the population had been vaccinated with at least one dose during the second and third time period, reducing the risk of contracting (symptomatic) COVID-19.

Thirdly, changes in societal / social behaviours may mean that contribution from occupational exposures on risk of infection is reduced compared to exposures outside the workplace. The omicron wave was more likely to lead to infections within the household (20).

In addition, the fact that fewer people went to their workplaces during time period 1, and people were likely to attend fewer social activities outside of work (21), the relative contribution of occupation to the risk of contracting COVID-19 was greater, which made the JEM perform better during this time period. Further developments of the COVID-19 JEM should include different periods reflecting different levels of ‘working from home’.

The reduced risk of infection in time periods 2 and 3 observed for the high risk group for the domain relating to the likelihood of contact with COVID-19 could be explained by the fact that this group mainly consists of health and social care workers. Previous surveys on the UK health sector suggested logistical issues with PPE in the UK health sector including a lack of means, inadequate training and inconsistent guidance during the first period of the pandemic (22). In addition, analysis amongst these occupational groups have shown lower risks in later stages of the pandemic, possibly related to better access to PPE, being a priority group for vaccination, or previous infection (23). Another exception is the domain relating to use of face coverings, where the low risk group appears to have a reduced risk of infection in time periods 2 and 3 when compared to the ‘no risk/work from home’ group; perhaps due to an increased risk in the homeworker group as restrictions changed.

In the Netherlands, the eight dimensions of the Dutch version of this JEM were recently validated against self-reported data related to the transmission risks and mitigation measures as well as COVID-19 infections within the past 12 months from the Netherlands Working Conditions Survey COVID-19 (NWCS-COVID-19) cohort study (12). Results showed good comparability between risk scores derived from the self-reported measures and the risks derived from the JEM. Self-reported infection data were collected during March 2021 and thereby the 12 months period that those cover is roughly comparable to our data covered by time period 1. The results of the Dutch study were very similar to the results of our study for the same period with higher JEM scores for the first six dimensions associated with a higher risk of having had COVID-19 compared with the reference score of “no risk”. In concordance with our findings, they also observed less strong associations for the dimensions “work location” and “face covering” and no evidence of association with the precarious work dimensions. Previous studies have suggested that migrant workers are at increased risk of SARS-CoV-2 infection at work due to their status as essential worker with often inequitable working and living conditions (24-26). Our results were imprecise due to small numbers in the higher risk categories so we cannot make conclusions. We did not find an association between a precarious occupation and SARS-CoV-2 but again there was a lot of uncertainty; earlier findings (5) suggested links between insecure employment and increased risk of COVID-19. The strict definition employed for precarious employment in the JEM was based on the proportion of workers with zero hours contracts. Precarity in employment is a complex issue characterised by employment insecurity, income inadequacy, and the lack of rights and protection and perhaps the JEM definition should be revised to capture precarity more broadly (27).

Several other studies assessed the association between various transmission and mitigation factors at work and the risk of COVID-19 (9, 28-31). Voko et al (31), evaluated the effect of social distancing on COVID-19 prevention in 28 European countries using incidence data from the European Centre for Disease Prevention and Control and index of social distancing developed from Google COVID-19 Community Mobility Reports. An increased social distance index was associated with fewer cases of infection on a daily basis (15). In regression analysis of data from the O*NET database, exposure to disease/infection in the workplace and a requirement of close physical proximity to other people during work singlehandedly explained more than 47% of disease prevalence variance (11). Another study examined the effect of ventilation, frequency of workplace contact and of the indoor/outdoor working environment contrast against serological SARS-CoV-2 status data from 3761 UK adult workers (12). Seropositivity was higher among workers with daily close contact, compared to those with intermediate-frequency contact and/or no work-related close contact. The risk of positive infection status was also generally elevated among workers in indoor trades, health care and in poor ventilated workplaces. The importance of ventilation by natural airflows was found in a study involving an outpatient building in Shenzen, China (14). Although not the main pathway for exposure to SARS-CoV-2, surface contamination has been suggested to still be important and more prevalent in frequently used, uncleaned, surfaces (29, 32, 33). These findings further support the applicability and relevance of a COVID-19 JEM for the assessment of infection/disease risk when individual data are unavailable, insufficient, or unfeasible to obtain. Some domains appear more relevant than others; focus on social distancing and indoor/outdoor working may capture the majority of variation while minimising collinearity.

Strengths of this study include the use of infection data from two independent cohorts, with a large number of participants including multiple time periods with different rates of infection as well as different restrictions and public health measures. One cohort (CIS) used repeated testing for all participants, hence frequency of testing was not related to occupation; in the other cohort (Virus Watch) testing on a subsample was used to check the reliability of the results relating to self-reported tests.

A JEM has limitations; differences exist within job codes which can dilute the results. We were unable to discriminate between infections acquired at work and those acquired outside work. The level of non-occupational exposure to SARS-CoV-2 was not included in analyses and will have changed substantially over time. In addition, it is possible that participants changed job (code) or lost their job during the study period, although for the stratified analyses by time period, occupation at the start of the time period was used. Finally, it is possible that infections between visits or prior to starting the study were missed. Missing data could be related to occupation, for example shift workers being unavailable at the time of the CIS study visits or too busy to respond to the Virus Watch questionnaire.

Overall, findings suggest that the COVID-19 JEM is a useful tool to assess occupational exposure to SARS-CoV-2 during the first time period, especially when more precise and/or individual level data are lacking. In order to extend this to later time periods, the addition of a dimension on vaccination and a job specific variable to indicate the likelihood of attending work during a certain time period should be considered. These adjustments could improve performance, resulting in a more accurate research instrument.

## Conclusions

We evaluated whether domains of a COVID-19 JEM were associated with the risk of SARS-CoV-2 infection. We observed clear exposure-response relationships in the early phase of the COVID-19 pandemic between the scores of the JEM relating to the number of contacts, the nature of contacts and social distancing, and the risk of SARS-CoV-2 infection. These relationships were accordant across the two cohorts involved and consistent with earlier results from a validation exercise of the JEM using Dutch data.(29). However, these observed relationships were not persistent over time. At the latter phases of the pandemic there was little evidence of a relationship between the domains of the JEM and SARS-CoV-2. Explanations could include i) a reduced role of workplace for infection risk as society opened, ii) changes in risk factors and effectiveness of control measures due to different variants and vaccinations introduction, iii) an increased potential for JEM misclassification amid changes in policy measures and restrictions. These findings suggest that the COVID-19 JEM is a useful tool for assessing risk of exposure to SARS-CoV-2 in the workplace during the period up to the end of the 2^nd^ wave of the pandemic. Modifications of the JEM may improve the performance of the JEM in the latter periods of the pandemic.

## Supporting information

STROBE checklist

## Data Availability

ONS CIS data can be accessed only by researchers who are Office of National Statistics (ONS) accredited researchers. Researchers can apply for accreditation through the Research Accreditation Service. Access is through the Secure Research Service (SRS) and approved on a project basis. For further details see https://www.ons.gov.uk/aboutus/whatwedo/statistics/requestingstatistics/approvedresearcherscheme.
Virus Watch aim to share aggregate data from this project on our website and via a "Findings so far" section on our website - https://ucl-virus-watch.net/. We also share some individual record level data on the Office of National Statistics Secure Research Service. In sharing the data we will work within the principles set out in the UKRI Guidance on best practice in the management of research data. Access to use of the data whilst research is being conducted will be managed by the Chief Investigators (ACH and RWA) in accordance with the principles set out in the UKRI guidance on best practice in the management of research data. We will put analysis code on publicly available repositories to enable their reuse.

## Declaration

This work was produced using statistical data from ONS. The use of the ONS statistical data in this work does not imply the endorsement of the ONS in relation to the interpretation or analysis of the statistical data. This work uses research datasets which may not exactly reproduce National Statistics aggregates.

## Ethics

The COVID-19 Infection Survey received ethical approval from the South Central Berkshire B Research Ethics Committee (20/SC/0195). All participants provided informed consent. For use of this data for this project statistics authority self-assessment classified the study as low risk. This assessment was approved by the Office for National Statistics (ONS) Research Accreditation Panel.

The Virus Watch study was approved by the Hampstead NHS Health Research Authority Ethics Committee: 20/HRA/2320, and conformed to the ethical standards set out in the Declaration of Helsinki. All participants provided informed consent for all aspects of the study.

## Data Availability

ONS CIS data can be accessed only by researchers who are Office of National Statistics (ONS) accredited researchers. Researchers can apply for accreditation through the Research Accreditation Service. Access is through the Secure Research Service (SRS) and approved on a project basis. For further details see https://www.ons.gov.uk/aboutus/whatwedo/statistics/requestingstatistics/approvedresearcherscheme.

Virus Watch aim to share aggregate data from this project on our website and via a “Findings so far” section on our website - https://ucl-virus-watch.net/. We also share some individual record level data on the Office of National Statistics Secure Research Service. In sharing the data we will work within the principles set out in the UKRI Guidance on best practice in the management of research data. Access to use of the data whilst research is being conducted will be managed by the Chief Investigators (ACH and RWA) in accordance with the principles set out in the UKRI guidance on best practice in the management of research data. We will put analysis code on publicly available repositories to enable their reuse.

## Supplementary Material

**S Table 1.** Odds ratios (95% CIs) by level of exposure for Domains 1-4 of the Job Exposure Matrix for two cohorts (CIS and Virus Watch). Models adjusted for age quintile, sex, ethnicity, IMD, region, household size, urban vs rural area, and presence of health conditions. N = 244,470 and N=16,479 for CIS and Virus Watch cohorts respectively.

|  | Domain 1<br>(Number) |  | Domain 2<br>(Nature) |  | Domain 3<br>(Surfaces) |  | Domain 4<br>(Location) |  |
| --- | --- | --- | --- | --- | --- | --- | --- | --- |
|  | CIS | Virus Watch | CIS | Virus Watch | CIS | Virus Watch | CIS | Virus Watch |
| No Risk | 1 | 1 | 1 | 1 | 1 | 1 | 1 | 1 |
| Low Risk | 1.07<br>(1.02,<br>1.12) | 0.99 (0.88,<br>1.11) | 1.10 (1.06,<br>1.15) | 1.13 (1.02,<br>1.25) | 1.13<br>(1.08,<br>1.19) | 1.19 (1.07,<br>1.32) | 1.14<br>(1.04,<br>1.24) | 1.08 (0.84,<br>1.37) |
| Elevated Risk | 1.11<br>(1.07,<br>1.16) | 1.22 (1.11,<br>1.35) | 1.17 (1.13,<br>1.22) | 1.17 (1.07,<br>1.27) | 1.08<br>(1.03,<br>1.14) | 1.17 (1.04,<br>1.30) | 1.20<br>(1.12,<br>1.27) | 1.03 (0.87,<br>1.22) |
| High Risk | 1.19<br>(1.14,<br>1.24) | 1.24 (1.12,<br>1.37) | 0.97 (0.90,<br>1.04) | 1.48 (1.26,<br>1.72) | 1.15<br>(1.11,<br>1.20) | 1.26 (1.15,<br>1.38) | 1.12<br>(1.08,<br>1.16) | 1.19 (1.10,<br>1.29) |
| Missing | 1.18<br>(1.14,<br>1.23) | 0.89 (0.70,<br>1.11) | 1.18 (1.14,<br>1.23) | 0.89 (0.71,<br>1.12) | 1.17<br>(1.13,<br>1.22) | 0.90 (0.72,<br>1.13) | 1.18<br>(1.14,<br>1.23) | 0.89 (0.70,<br>1.11) |

**S Table 2.** Odds ratios (95% CIs) by level of exposure for Domains 5-8 of the Job Exposure Matrix for two cohorts (CIS and Virus Watch). Models adjusted for age quintile, sex, ethnicity, IMD, region, household size, urban vs rural area, and presence of health conditions. Note that Low Risk is used as the reference category for Domain 7 for the Virus Watch cohort, due to small numbers in the No Risk category. N = 244,470 and N=16,479 for CIS and Virus Watch cohorts respectively.

|  | Domain 5<br>(Distancing) |  | Domain 6<br>(Face covering) |  | Domain 7<br>(Insecurity) |  | Domain 8<br>(Migrants) |  |
| --- | --- | --- | --- | --- | --- | --- | --- | --- |
|  | CIS | Virus Watch | CIS | Virus Watch | CIS | Virus Watch | CIS | Virus Watch |
| No Risk | 1 | 1 | 1 | 1 | 1 | 1.10 (0.35,<br>2.86) | 1 | 1 |
| Low Risk | 1.06 (1.02,<br>1.10) | 1.04 (0.95,<br>1.14) | 1.07 (1.02,<br>1.12) | 1.29 (1.16,<br>1.44) | 0.83 (0.55,<br>1.26) | 1 | 0.98 (0.95,<br>1.01) | 1.00 (0.93,<br>1.07) |
| Elevated Risk | 1.15 (1.11,<br>1.21) | 1.22 (1.11,<br>1.35) | 1.15 (1.11,<br>1.20) | 1.12 (1.03,<br>1.22) | 0.80 (0.53,<br>1.21) | 1.07 (0.99,<br>1.15) | 1.09 (0.99,<br>1.19) | 1.56 (1.23,<br>1.96) |
| High Risk | 1.28 (1.22,<br>1.35) | 1.54 (1.36,<br>1.74) | 1.05 (0.90,<br>1.23) | 1.47 (1.11,<br>1.93) | 1.00 (0.63,<br>1.58) | 1.38 (0.76,<br>2.40) | 1.42 (1.19,<br>1.69) | 0.93 (0.52,<br>1.59) |
| Missing | 1.18 (1.14,<br>1.23) | 0.89 (0.70,<br>1.11) | 1.18 (1.14,<br>1.23) | 0.89 (0.70,<br>1.11) | 0.89 (0.59,<br>1.35) | 0.83 (0.66,<br>1.03) | 1.08 (1.05,<br>1.12) | 0.81 (0.64,<br>1.01) |

**S Table 3.**
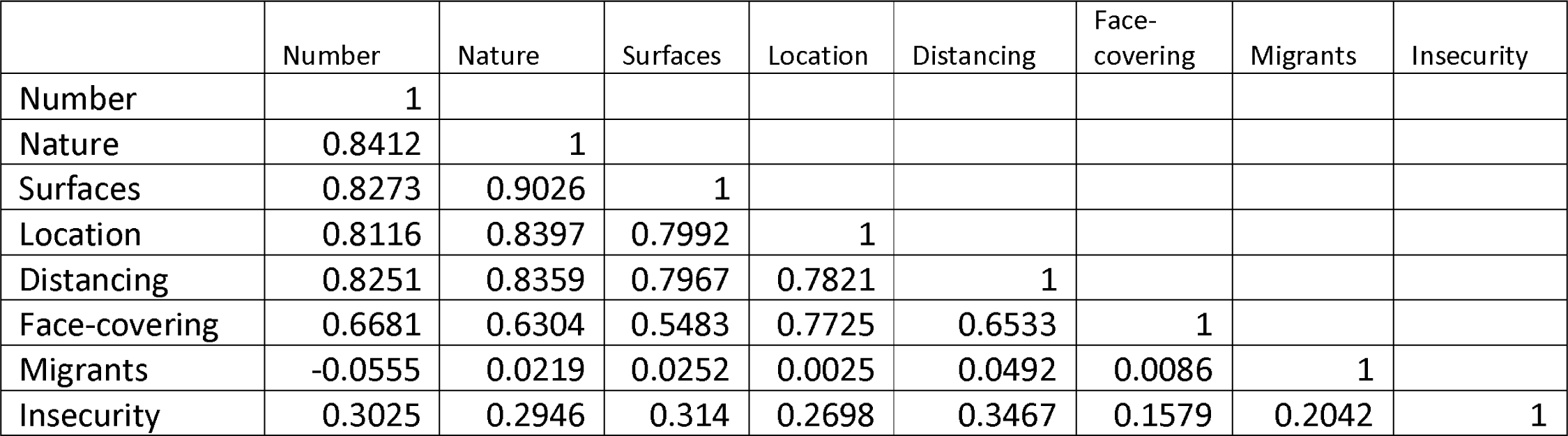
Spearman’s rank correlation matrix for 8 domains of the JEM, using first available JEM code. N=176 713 participants with available SOC in the CIS

|  | Number | Nature | Surfaces | Location | Distancing | Face-covering | Migrants | Insecurity |
| --- | --- | --- | --- | --- | --- | --- | --- | --- |
| Number | 1 |  |  |  |  |  |  |  |
| Nature | 0.8412 | 1 |  |  |  |  |  |  |
| Surfaces | 0.8273 | 0.9026 | 1 |  |  |  |  |  |
| Location | 0.8116 | 0.8397 | 0.7992 | 1 |  |  |  |  |
| Distancing | 0.8251 | 0.8359 | 0.7967 | 0.7821 | 1 |  |  |  |
| Face-covering | 0.6681 | 0.6304 | 0.5483 | 0.7725 | 0.6533 | 1 |  |  |
| Migrants | -0.0555 | 0.0219 | 0.0252 | 0.0025 | 0.0492 | 0.0086 | 1 |  |
| Insecurity | 0.3025 | 0.2946 | 0.314 | 0.2698 | 0.3467 | 0.1579 | 0.2042 | 1 |

## References

1. Taylor L. Covid-19: True global death toll from pandemic is almost 15 million, says WHO. BMJ. 2022;377:o1144.

2. Mutambudzi M, Niedzwiedz C, Macdonald EB, Leyland A, Mair F, Anderson J, et al. Occupation and risk of severe COVID-19: prospective cohort study of 120 075 UK Biobank participants. Occupational and Environmental Medicine. 2021;78(5):307.

3. Nafilyan V, Pawelek P, Ayoubkhani D, Rhodes S, Pembrey L, Matz M, et al. Occupation and COVID-19 mortality in England: a national linked data study of 14.3 million adults. Occupational and Environmental Medicine. 2021:oemed-2021-107818.

4. Oude Hengel KM, Burdorf A, Pronk A, Schlünssen V, Stokholm ZA, Kolstad HA, et al. Exposure to a SARS-CoV-2 infection at work: development of an international job exposure matrix (COVID-19-JEM). Scandinavian journal of work, environment & health. 2022(1):61–70.

5. Burdorf A, Porru F, Rugulies R. The COVID-19 pandemic: one year later - an occupational perspective. Scandinavian journal of work, environment & health. 2021;47(4):245–7.

6. Nelson KN, Siegler AJ, Sullivan PS, Bradley H, Hall E, Luisi N, et al. Nationally Representative Social Contact Patterns among U.S. adults, August 2020-April 2021. medRxiv. 2022:2021.09.22.21263904.

7. Beale S, Hoskins S, Byrne T, Fong WLE, Fragaszy E, Geismar C, et al. Workplace contact patterns in England during the COVID-19 pandemic: Analysis of the Virus Watch prospective cohort study. The Lancet Regional Health - Europe. 2022;16:100352.

8. Baker MG, Peckham TK, Seixas NS. Estimating the burden of United States workers exposed to infection or disease: A key factor in containing risk of COVID-19 infection. PLOS ONE. 2020;15(4):e0232452.

9. Beale S, Patel P, Rodger A, Braithwaite I, Byrne T, Fong WLE, et al. Occupation, work-related contact and SARS-CoV-2 anti-nucleocapsid serological status: findings from the Virus Watch prospective cohort study. Occupational and Environmental Medicine. 2022:oemed-2021-107920.

10. Chen Y-H, Glymour M, Riley A, Balmes J, Duchowny K, Harrison R, et al. Excess mortality associated with the COVID-19 pandemic among Californians 18–65 years of age, by occupational sector and occupation: March through November 2020. PLOS ONE. 2021;16(6):e0252454.

11. Billingsley S, Brandén M, Aradhya S, Drefahl S, Andersson G, Mussino E. COVID-19 mortality across occupations and secondary risks for elderly individuals in the household: A population register-based study. Scandinavian journal of work, environment & health.

12. van der Feltz S, Peters S, Pronk A, Schlünssen V, Stokholm ZA, Kolstad HA, et al. Validation of a COVID-19 Job Exposure Matrix (COVID-19-JEM) for Occupational Risk of a SARS-CoV-2 Infection at Work: Using Data of Dutch Workers. Ann Work Expo Health. 2022.

13. Office IL. International Standard Classification of Occupations Geneva 2008 [Available from: https://www.ilo.org/public/english/bureau/stat/isco/docs/publication08.pdf.

14. ONS. SOC2010 volume 2: the structure and coding index 2010 [Available from: https://www.ons.gov.uk/methodology/classificationsandstandards/standardoccupationalclassificationsoc/soc2010/soc2010volume2thestructureandcodingindex.

15. Statistics OoN. Classifying the Standard Occupational Classification 2020 (SOC 2020) to the International Standard Classification of Occupations (ISCO-08). UK 2020.

16. ONS. Annual population survey (APS) QMI 2012 [Available from: https://www.ons.gov.uk/employmentandlabourmarket/peopleinwork/employmentandemployeetypes/methodologies/annualpopulationsurveyapsqmi.

17. ONS. Labour Force Survey (LFS) QMI 2015 [Available from: https://www.ons.gov.uk/employmentandlabourmarket/peopleinwork/employmentandemployeetypes/methodologies/labourforcesurveylfsqmi.

18. StataCorp. Stata Statistical Software: Release 17. College Station, TX: StataCorp LLC; 2021.

19. Greenhalgh T, Jimenez JL, Prather KA, Tufekci Z, Fisman D, Schooley R. Ten scientific reasons in support of airborne transmission of SARS-CoV-2. The Lancet. 2021;397(10285):1603–5.

20. Jalali N, Brustad HK, Frigessi A, MacDonald E, Meijerink H, Feruglio S, et al. Increased household transmission and immune escape of the SARS-CoV-2 Omicron variant compared to the Delta variant: evidence from Norwegian contact tracing and vaccination data. medRxiv. 2022:2022.02.07.22270437.

21. J P. Homeworking and spending during the coronavirus (COVID-19) pandemic, Great Britain: April 2020 to January 2022. In: Statistics OoN, editor. 2022.

22. Mantelakis A, Spiers HVM, Lee CW, Chambers A, Joshi A. Availability of Personal Protective Equipment in NHS Hospitals During COVID-19: A National Survey. Ann Work Expo Health. 2021;65(1):136–40.

23. Rhodes S, Wilkinson J, Pearce N, Mueller W, Cherrie M, Stocking K, et al. Occupational differences in SARS-CoV-2 infection: analysis of the UK ONS COVID-19 infection survey. Journal of Epidemiology and Community Health. 2022:jech-2022-219101.

24. Reid A, Ronda-Perez E, Schenker MB. Migrant workers, essential work, and COVID-19. Am J Ind Med. 2021;64(2):73–7.

25. Burton-Jeangros C, Duvoisin A, Lachat S, Consoli L, Fakhoury J, Jackson Y. The Impact of the Covid-19 Pandemic and the Lockdown on the Health and Living Conditions of Undocumented Migrants and Migrants Undergoing Legal Status Regularization. Frontiers in Public Health. 2020;8.

26. Liem A, Wang C, Wariyanti Y, Latkin CA, Hall BJ. The neglected health of international migrant workers in the COVID-19 epidemic. The Lancet Psychiatry. 2020;7(4):e20.

27. Kreshpaj B, Orellana C, Burström B, Davis L, Hemmingsson T, Johansson G, et al. What is precarious employment? A systematic review of definitions and operationalizations from quantitative and qualitative studies. Scandinavian journal of work, environment & health. 2020;46(3):235–47.

28. Zhang M. Estimation of differential occupational risk of COVID-19 by comparing risk factors with case data by occupational group. Am J Ind Med. 2021;64(1):39–47.

29. Kampf G, Todt D, Pfaender S, Steinmann E. Persistence of coronaviruses on inanimate surfaces and their inactivation with biocidal agents. Journal of Hospital Infection. 2020;104(3):246–51.

30. Li C, Tang H. Study on ventilation rates and assessment of infection risks of COVID-19 in an outpatient building. Journal of Building Engineering. 2021;42:103090-.

31. Vokó Z, Pitter JG. The effect of social distance measures on COVID-19 epidemics in Europe: an interrupted time series analysis. GeroScience. 2020;42(4):1075–82.

32. Hennessy NT, Toomey S, Gautier V, O’Reilly S, de Barra E, Hanrahan EO, et al. COVID-19 contamination of high-touch surfaces in the public domain. Irish journal of medical science. 2022:1–2.

33. Silva PGD, Gonçalves J, Lopes AIB, Esteves NA, Bamba GEE, Nascimento MSJ, et al. Evidence of Air and Surface Contamination with SARS-CoV-2 in a Major Hospital in Portugal. Int J Environ Res Public Health. 2022;19(1).

